# Repetitive Transcranial Magnetic Stimulation for Tobacco Treatment in Cancer Patients: A Preliminary Report of a One-Week Treatment

**DOI:** 10.1101/2022.03.11.22269298

**Authors:** Xingbao Li, Benjamin A. Toll, Matthew J. Carpenter, Paul J Nietert, Morgan Dancy, Mark S. George

## Abstract

**Background:** Smoking cessation represents a significant opportunity to improve cancer survival rates, reduce the risk of cancer treatment complications, and improve quality of life. However, about half of cancer patients who smoke continue to smoke despite the availability of several treatments. Previous studies demonstrate that repetitive transcranial magnetic stimulation (rTMS) over the left dorsolateral prefrontal cortex (DLPFC) decreases cue craving, reduces cigarette consumption, and increases the quit rate in tobacco use disorder. We investigated whether 5 sessions of rTMS can be safely and efficaciously used for smoking cessation in cancer patients.

**Methods:** We enrolled 11 treatment-seeking smokers with cancer (> 5 cigarettes per day) in a randomized, double-blind, sham-controlled proof-of-concept study. Participants received 5 daily sessions of active 10Hz rTMS of the left DLPFC (3000 pulses per session) or sham rTMS, and were followed up for 1 month via phone assessments. Main outcomes included reductions in the number of smoked-cigarettes per day (primary) and craving (secondary). Adverse effects were reported daily by participants.

**Results:** Seven of 11 participants completed 5 sessions of rTMS over one week. Compared to sham treatment (n = 4), the active rTMS (n = 3) exhibited modest effects overtime on smoking (Cohen’s *f*^*2*^ effect size of 0.16) and large effects on cue craving (Cohen’s *f*^*2*^ = 0.40). No serious side effects related to rTMS were reported in the treatment.

**Conclusions:** Five sessions of daily rTMS over the left DLPFC might benefit cancer patients who smoke cigarettes. However, further evidence is needed to determine with more certainty its therapeutic effect and adverse effects for cancer patients who smoke cigarettes.

## Introduction

The majority of smokers diagnosed with cancer continue to smoke after diagnosis, even in the context of an intention to quit and attempts to do so ^1^. Previous observational studies have reported an estimated smoking prevalence of 45-60% among patients at the time of cancer diagnosis ^2^. Furthermore, about 47-60% of patients with newly diagnosed cancer continue to smoke, ^3,4^ and relapse rates ranging from 50% to 83% have been reported among cancer survivors^5^. Continued smoking after a cancer diagnosis increases the risk of second primary tumors and cancer recurrence is a cause of treatment complications and decreases survival ^6,7.*8*^

The US National Comprehensive Cancer Network (NCCN) published practice guidelines emphasizing combined behavioral and medication treatment to bolster cessation outcomes among this population ^9 10^. To date, there are different and complementary approaches to quitting smoking. In general, the most effective way to quit involves a combination of counseling and medications ^11^; however, given modest quit rates with existing treatments, more novel methods to improve smoking cessation among cancer patients are needed^8^.

Transcranial magnetic stimulation (TMS) is a noninvasive brain stimulation technique that can focally stimulate the brain of an awake individual ^12^. Recently, the U.S. Food and Drug Administration (FDA) approved high frequency (HF)-repetitive TMS (rTMS) (Brainsway) for smoking cessation for adult smokers ^13^. Previously, our double-blind sham-controlled, randomized clinical trial showed that 2-week imaged-guided rTMS of the left dorsal lateral prefrontal cortex (DLPFC) reduced cigarette consumption and increased quit rates ^14^. Overall, these previous studies demonstrate that HF-rTMS of the DLPFC can attenuate cigarette consumption, ^14-16^ craving, ^14,15,17^ and increase quit rates ^14,16^. However, to date, no study has been reported to use rTMS for tobacco treatment for cancer patients. We investigated whether 5 sessions of rTMS was a feasible and safe novel method of tobacco treatment for cancer patients, and we assessed the effect sizes for rTMS as compared to sham treatment, which can be useful (but not sufficient) for hypothesis generation^18^.

## MATERIALS and METHODS

### Study Design

This randomized, double-blind, sham-controlled proof of concept study was conducted at the Medical University of South Carolina (MUSC) in Charleston, South Carolina, USA. The study consisted of 5 daily rTMS sessions over the left DLPFC across 1 week, with a 1-month follow-up by phone. Outcome measures included the following: Self-reported cigarette consumption, cue-induced craving measures, withdrawal symptoms, and carbon monoxide (CO) level.

The Institutional Review Board at MUSC approved all study procedures, and the study was registered on ClinicalTrials.gov (NCT02401672).

### Participants

To be eligible for this study, the patient must have met all of the following criteria: (1) Completed cancer surgery treatment ≥ 6 months or other current cancer therapeutics; (2) Have been diagnosed with early-stage lung cancer, breast cancer or prostate cancer; (3) Smoke 5+ cigarettes per day (CPD); (4) Show symptoms of nicotine dependence as determined by the Fagerstrom Test for Nicotine Dependence (FTND) ≥1; (5) Be able to comply with protocol requirements and likely to complete all study procedures; (6) Be willing to consider quitting smoking; (7) Have no active cardiac, neurologic, or psychiatric illness. Exclusion criteria included: (1) Current dependence, defined by DSM-V criteria, on any psychoactive substances other than nicotine or caffeine; (2) Contraindication to rTMS (history of neurological disorder or seizure, increased intracranial pressure, brain surgery, or head trauma with loss of consciousness for > 15 minutes, implanted electronic device, metal in the head, or pregnancy); (3) History of autoimmune, endocrine, viral, or vascular disorder affecting the brain; (4) Use of other tobacco treatments at the time of the study procedures.

### Randomization and Blinding

Eligible participants were randomized in a 1:1 manner to either active 10Hz rTMS of the left DLPFC (3000 pulses per session) or sham rTMS. A unique participant randomization code was assigned prior to the first rTMS session. A staff member uninvolved in treatment delivery or rating scale administration selected the rTMS coil (Active/Sham) for each participant.

### TMS Procedures

Participants were instructed to abstain from smoking for at least 2 hours before each treatment, intended to increase the degree of craving during the treatment as per previous protocols ^17,19^. Participants were asked to reduce the number of cigarettes smoked.

#### rTMS Therapy Procedures

These procedures were based on a modified version of the NeuroStar XPLOR Clinical Research System (Neuronetics, Inc), which was used in our previous study ^14^. At entry, we determined each subject’s resting motor threshold (rMT); all rTMS dosings were given relative to this value. The iron-core, solid-state figure-of-8 coil was positioned over the area of the skull roughly corresponding to the motor cortex and then systematically moved and adjusted until each pulse resulted in the isolated movement of the right thumb. rMT was determined with the Neurostar algorithm, which provides an iterated estimate of the rMT^20,21^.

After the patient’s rMT was established, the TMS coil was moved to the anterior direction by 5.5 cm. To ensure that patients received the right dose to the right location every time, we used contact sensors for each treatment with a rotation point about the tip of the subject’s nose. At a visit, after providing informed consent, participants were fitted with a white lycra swim cap. This cap was worn during all TMS sessions in order to ensure proper placement of the TMS coil across visits.

Active rTMS was administered at 100% rMT, at 10 Hz for 5-second trains, with an inter-train interval of 10 seconds. Treatment sessions lasted for 15 minutes (60 trains) with 3000 pulses/session. Sham rTMS was delivered via a procedure used previously ^22,23^, in which the sensation of active rTMS is mimicked using time-locked electrical stimulation at the target treatment site without a magnetic intervention. Both Active and Sham rTMS sessions were scheduled daily for 5 consecutive weekdays. Treatment was not administered on the weekend.

#### Cue Provocation

We used structured 1.5 min exposure and interactions with real-life smoking paraphernalia (cigarettes, ashtray, lighter) ^14^ immediately before each rTMS session. While rTMS was administered, participants watched a 15-minute video displaying smoking cues^17^ (scenes of individuals smoking in various environments) displayed on an iPad placed on a tripod at the foot of the treatment chair.

### Evaluation of Cigarette Consumption, Cued Craving, and Biomarkers

The number of cigarettes smoked per day (printed pre-prepared cigarette diary brought to the TMS lab for each session) ^24^ was evaluated by subjective self-report.

The Questionnaire of Smoking Urges-Brief (QSU-B) ^25^ was assessed before and after each TMS session. CO levels were measured before each TMS treatment using the Micro Smokerlyzer Breath Carbon Monoxide Monitor.

### Treatment adherence, Visit attendance, and Adverse Events

Adverse events were recorded over the 5-day treatment course. Anticipated events include pain or discomfort on the site of rTMS, headache, muscle twitching, back pain, anxiety, and insomnia.

### Statistical Analysis

Descriptive statistics were used to summarize outcomes by treatment group. Outcome measures were then compared between treatment groups Mixed-Models for Repeated Measures (MMRM). While p-values from the MMRM are reported, our primary focus was the estimation of effect sizes since this was a pilot study and largely hypothesis-generating. Cohen’s *f*^*2*^ effect sizes ^26^ were calculated based on results of the MMRMs. The MMRMs included fixed effects for treatment group (sham vs. active), session number (1 through 5), treatment group x session number interaction, the baseline value of the outcome, and within-session timing (pre-vs. post-TMS), if applicable. We accounted for within-subject clustering using an AR(1) or compound symmetry (CS) error structure, depending on model convergence status and which structure yielded the lowest AIC value. Descriptive statistics were calculated using IBM SPSS Statistics 25 (IBM, Endicott, New York), and the MMRMs were constructed using SAS v9.4 (SAS Institute, Cary, NC). Model construction details are provided in the Supplementary Material.

## RESULTS

### Participant Characteristics

Twenty individual smokers who were cancer patients were screened by phone, of which 13 were invited for a screening visit, 11 met study criteria, all of whom enrolled. Three participants declined before treatment initiation. Eight participants were randomized and started the treatment. Among the 8 participants who started treatment, 4 received sham treatment, and the other 4 received active rTMS treatment. Seven (87.5%) completed all 5 sessions of rTMS. One who received active treatment dropped out of the study at the 3^rd^ session visit because of a serious adverse event (SAE). Participant demographics and baseline smoking-related variables are summarized in Table 1.

**Table 1:** Demographic Information (Baseline)

|  | **Sham rTMS** (n = 4) | **Active rTMS** (n = 3) | **P-Value** |
| --- | --- | --- | --- |
| Age, mean (SD) | 64.5 (3.4) | 62.7 (3.1) | 0.49 |
| Gender, M/F | 4/0 | 2/1 | 0.21 |
| Cancer, Lung/others | 2/2 | 2/1 | 0.65 |
| Years smoked, mean (SD) | 41.8 (11.3) | 49.7 (4.6) | 0.34 |
| Smoking Start Age, Mean (SD) | 20.0 (6.9) | 12.7 (2.1) | 0.14 |
| Cigarettes per day, mean (SD) | 11.2 (5.9) | 17.0 (12.1) | 0.43 |
| FTND score, mean (SD) | 4.3 (3.9) | 5.7 (3.5) | 0.64 |

FTND: Fagerstrom Test for Nicotine Dependence

### Self-reported Cigarette Consumption

Baseline cigarettes smoked per day (CPD) were comparable between study groups; however, after 5 TMS sessions, the mean number of CPDs was moderately lower among the active TMS group than the sham group, resulting in a Cohen’s *f*^*2*^ effect size of 0.16, corresponding to a medium effect size^27^ (sham: baseline mean [SD] = 12.0 [5.4]; 5^th^ session: 10.8 [6.7]; Active: baseline: 17.0 [12.1]; 5^th^ session: 10.0 [6.2]; *p*=0.14). (Figure 1. A)

**Figure 1.**
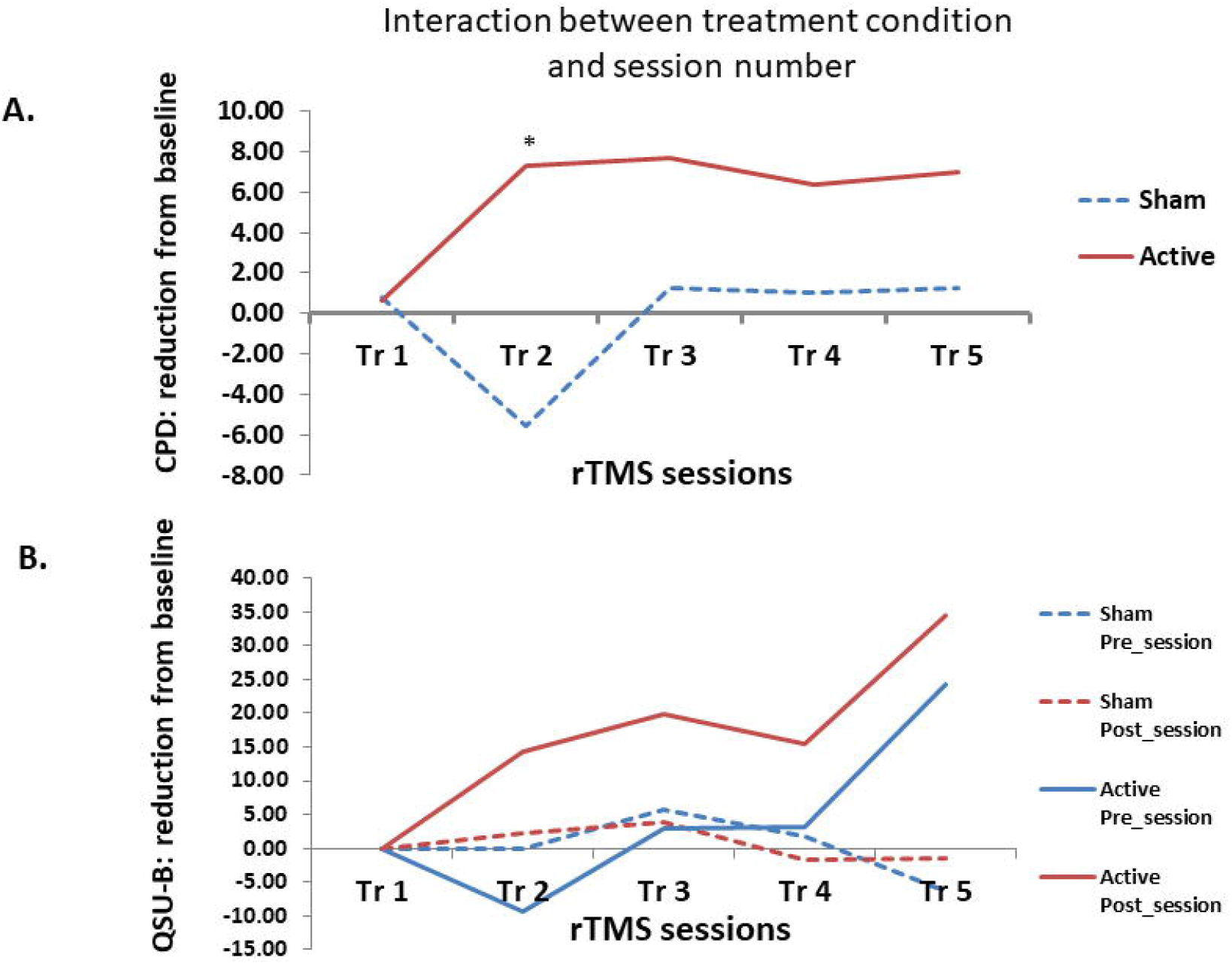
One-week rTMS reduces cigarette consumption and cue-craving in cancer patients A. rTMS reduced CPD to a significantly greater degree than sham rTMS (p <0.005). (Sham: blue, Active: red) Post hoc, there is a difference between the two treatments at 2^nd^ visit, * p < 0.05 B. rTMS reduced cue-craving to a significantly greater degree than sham rTMS (p <0.0001). (Sham: blue, Active: red; Pre-session measure: solid, Post-session measure: dahes)

### Subjective Cued Craving

#### Questionnaire of Smoking Urges-Brief (QSU-B)

Results indicated a large difference between groups at the end of session 5 (Cohen’s *f*^*2*^ = 0.40). The model showed a significant (*p*=0.001) interaction between treatment group and session number, indicating that the mean QSU-B scores were relatively unchanged over the 5 sessions in the sham TMS group (mean [SD] at beginning of session 1: 36.0 [43.7] vs. end of session 5: 32.8 [44.7]) but markedly changed (lower over time) in the active TMS group (beginning of session 1: 45.3 [24.9] vs. end of session 5: 25.6 [24.1]) (Figure 1.B).

### Biochemical Measures CO levels

The mixed model analysis indicated a slight difference in CO levels between groups at the end of session 5, with CO levels slightly lower among subjects in the active group compared with the sham group (sham, session 1: mean [SD] = 11.8 [9.1] ppm, session 5: 13.0 [11.0]; Active, session 1: 7.3 [3.2], session 5: 6.7 [5.5], Cohen’s *f*^*2*^ = 0.02).

### Treatment Adherence, Visit Attendance, and Adverse Events

In general, rTMS was found to be safe for tobacco treatment with cancer patients. One participant reported back pain after sham treatment and recovered without treatment. One participant reported a metallic taste after active TMS. No other adverse events were reported. One of active treatment group had SAE at the 3^rd^ session visit. When the participant met the research staff for the 3^rd^ treatment, The PI (Li) found that the participant was a confusional state, disorientation, and difficulty following commands. The PI detected the SAE during the pre-session assessment and was thus deemed not to be related to TMS treatment.

## Discussion

The findings from this sham-controlled pilot trial suggest, but do not definitively prove, that five sessions of active rTMS over the left DLPFC can be feasibly and safely used for smoking cessationin cancer patients. The 10 Hz rTMS at 100% motor threshold rTMS protocol was well tolerated among participants, with few treatment-related adverse events. Feasibility is also supported in that all but one participant who initiated treatment completed all sessions. The treatment demands of rTMS may make it a challenge for all cancer patients, but our data show that those who start are receptive to finish. We found that 5 sessions of 10 Hz rTMS over the LDLPFC had a modest impact on cigarette smoking (CPD) and cue-induced craving (QSU-B) in cancer patients who smoke cigarettes, without serious side effects, although it is important to note that all of these findings would need to be confirmed in a larger definitive study. Results of this study are consistent with what we have found in healthy smokers ^14,17^.

Tobacco treatment for cancer patients who smoke should be a top priority for all physicians and healthcare providers ^28,29^. Daily smoking promotes tumor progression, increases the risk of second cancers, and decreases survival. Smoking increases side effects of chemotherapy and surgery and reduces the effectiveness of radiotherapy and chemotherapy; however, some cancer patients may be resistant to treatment and unmotivated to seek treatment to reduce or quit smoking. Therefore, a future larger study will be needed to determine whether multiple sessions of rTMS are efficacious in reducing cigarette consumption and/or increasing the quit rate among cancer patients who smoke.

## Supporting information

The results of other measures and the methods of modelling

## Data Availability

All data produced in the present study are available upon reasonable request to the authors

## Acknowledgments and COIs

This project was supported by the National Center for Advancing Translational Sciences of the National Institutes of Health under Grant Number UL1 TR001450 (South Carolina Clinical & Translational Research Institute discovery grant 1711). The work was also supported, in part, by NIH R21DA036752 (X Li) and NIH UG3 DA048507 (X Li)

Drs. Carpenter and Toll have served on Advisory Boards for Pfizer, and Dr. Toll testifies on behalf of plaintiffs who have filed litigation against the tobacco industry.

The other authors declare no conflicts of interest.

