## Supplementary material for "Repetitive Transcranial Magnetic Stimulation for Tobacco Treatment in Cancer Patients: A Preliminary Report of a One-Week Treatment": The results of other measures and the methods of modelling: Supplementary Material 03112022.docx

1. **Minnesota Nicotine Withdrawal Symptoms (MNWS)**

No meaningful differences between active and sham groups were noted in MNWS scores at session 1 (mean ± SD in sham group: 14.3 ± 8.7 vs. active group: 15.3 ± 6.4, p=0.87) or at session 5 (sham: 13.0 ± 6.3 vs. active: 10 ± 11.3, p=0.65), nor were there differences between groups in the change in MNWS scores (sham: -1.3 ± 4.6 vs. active: -5.3 ± 10.5, p=0.51).

2. **Mixed-Models for Repeated Measures Analysis**

**Note: The original data is available upon request from the corresponding author**.

Self-reported Cigarette Consumption

Note: The “CPDdata2” dataset is in long format, with a baseline measure and measurement during each of 5 rTMS sessions.

SAS programming code:

/* full model */

proc mixed data= CPDdata2;

class subject_ visit_num TrGroup;

model CigarettePerday = TrGroup visit_num TrGroup*visit_num CigarettePerday_bl / solution;

repeated / subject=subject_ type=ar(1) rcorr;

run;

/* null model - for aid in calculation of f squared statistic */

proc mixed data= CPDdata2;

class subject_ visit_num TrGroup;

model CigarettePerday = visit_num CigarettePerday_bl;

repeated / subject=subject_ type=ar(1) rcorr;

run;

Output from model estimation for the “full model”:

```
                                       The Mixed Procedure

                                        Model Information

                      Data Set                     WORK.CPDDATA2
                      Dependent Variable           CigarettePerday
                      Covariance Structure         Autoregressive
                      Subject Effect               Subject_
                      Estimation Method            REML
                      Residual Variance Method     Profile
                      Fixed Effects SE Method      Model-Based
                      Degrees of Freedom Method    Between-Within


                                     Class Level Information

                      Class        Levels    Values

                      Subject_          7    10002 10003 10004 10005 20001
                                             20002 20003
                      visit_num         6    0 1 2 3 4 5
                      TrGroup           2    X Y


                                           Dimensions

                               Covariance Parameters             2
                               Columns in X                     22
                               Columns in Z                      0
                               Subjects                          7
                               Max Obs per Subject               6


                                     Number of Observations

                           Number of Observations Read              42
                           Number of Observations Used              42
                           Number of Observations Not Used           0


                                        Iteration History

                   Iteration    Evaluations    -2 Res Log Like       Criterion

                           0              1       181.14762353
                           1              2       173.52687218      0.00000180
                           2              1       173.52676361      0.00000000


                                    Convergence criteria met.


                                       The Mixed Procedure

                         Estimated R Correlation Matrix for Subject_ 10002

            Row        Col1        Col2        Col3        Col4        Col5        Col6

              1      1.0000      0.4989      0.2489      0.1242     0.06196     0.03091
              2      0.4989      1.0000      0.4989      0.2489      0.1242     0.06196
              3      0.2489      0.4989      1.0000      0.4989      0.2489      0.1242
              4      0.1242      0.2489      0.4989      1.0000      0.4989      0.2489
              5     0.06196      0.1242      0.2489      0.4989      1.0000      0.4989
              6     0.03091     0.06196      0.1242      0.2489      0.4989      1.0000


                                 Covariance Parameter Estimates

                                Cov Parm     Subject     Estimate

                                AR(1)        Subject_      0.4989
                                Residual                  14.0068


                                         Fit Statistics

                              -2 Res Log Likelihood           173.5
                              AIC (Smaller is Better)         177.5
                              AICC (Smaller is Better)        178.0
                              BIC (Smaller is Better)         177.4


                                 Null Model Likelihood Ratio Test

                                   DF    Chi-Square      Pr > ChiSq

                                    1          7.62          0.0058


                                    Solution for Fixed Effects

                                                                        Standard
 Effect               X = sham, Y = active       visit_num   Estimate      Error     DF   t Value

 Intercept                                                    -2.6318     2.9386      4     -0.90
 TrGroup              X                                        4.4652     2.9178      4      1.53


                                    Solution for Fixed Effects

               Effect               X = sham, Y = active       visit_num   Pr > |t|

               Intercept                                                     0.4211
               TrGroup              X                                        0.2007


                                       The Mixed Procedure

                                    Solution for Fixed Effects

                                                                        Standard
 Effect               X = sham, Y = active       visit_num   Estimate      Error     DF   t Value

 TrGroup              Y                                             0          .      .       .
 visit_num                                       0             7.0000     3.0082     25      2.33
 visit_num                                       1             6.3333     2.9596     25      2.14
 visit_num                                       2            -0.3333     2.8598     25     -0.12
 visit_num                                       3            -0.6667     2.6483     25     -0.25
 visit_num                                       4             0.6667     2.1631     25      0.31
 visit_num                                       5                  0          .      .       .
 visit_num*TrGroup    X                          0            -5.7500     3.9795     25     -1.44
 visit_num*TrGroup    Y                          0                  0          .      .       .
 visit_num*TrGroup    X                          1            -5.8333     3.9152     25     -1.49
 visit_num*TrGroup    Y                          1                  0          .      .       .
 visit_num*TrGroup    X                          2             1.5833     3.7831     25      0.42
 visit_num*TrGroup    Y                          2                  0          .      .       .
 visit_num*TrGroup    X                          3             0.6667     3.5034     25      0.19
 visit_num*TrGroup    Y                          3                  0          .      .       .
 visit_num*TrGroup    X                          4            -0.4167     2.8615     25     -0.15
 visit_num*TrGroup    Y                          4                  0          .      .       .
 visit_num*TrGroup    X                          5                  0          .      .       .
 visit_num*TrGroup    Y                          5                  0          .      .       .
 CigarettePerday_bl                                            0.7430     0.1172      4      6.34


                                    Solution for Fixed Effects

               Effect               X = sham, Y = active       visit_num   Pr > |t|

               TrGroup              Y                                         .
               visit_num                                       0             0.0284
               visit_num                                       1             0.0423
               visit_num                                       2             0.9081
               visit_num                                       3             0.8033
               visit_num                                       4             0.7605
               visit_num                                       5              .
               visit_num*TrGroup    X                          0             0.1609
               visit_num*TrGroup    Y                          0              .
               visit_num*TrGroup    X                          1             0.1488
               visit_num*TrGroup    Y                          1              .
               visit_num*TrGroup    X                          2             0.6791
               visit_num*TrGroup    Y                          2              .
               visit_num*TrGroup    X                          3             0.8506
               visit_num*TrGroup    Y                          3              .
               visit_num*TrGroup    X                          4             0.8854
               visit_num*TrGroup    Y                          4              .
               visit_num*TrGroup    X                          5              .
               visit_num*TrGroup    Y                          5              .
               CigarettePerday_bl                                            0.0032


                                       The Mixed Procedure

                                  Type 3 Tests of Fixed Effects

                                            Num     Den
                     Effect                  DF      DF    F Value    Pr > F

                     TrGroup                  1       4       2.29    0.2051
                     visit_num                5      25       1.60    0.1980
                     visit_num*TrGroup        5      25       1.53    0.2168
                     CigarettePerday_bl       1       4      40.23    0.0032
```

Output from model estimation for the “null model”:

```
                                       The Mixed Procedure

                                        Model Information

                      Data Set                     WORK.CPDDATA2
                      Dependent Variable           CigarettePerday
                      Covariance Structure         Autoregressive
                      Subject Effect               Subject_
                      Estimation Method            REML
                      Residual Variance Method     Profile
                      Fixed Effects SE Method      Model-Based
                      Degrees of Freedom Method    Between-Within


                                     Class Level Information

                      Class        Levels    Values

                      Subject_          7    10002 10003 10004 10005 20001
                                             20002 20003
                      visit_num         6    0 1 2 3 4 5
                      TrGroup           2    X Y


                                           Dimensions

                               Covariance Parameters             2
                               Columns in X                      8
                               Columns in Z                      0
                               Subjects                          7
                               Max Obs per Subject               6


                                     Number of Observations

                           Number of Observations Read              42
                           Number of Observations Used              42
                           Number of Observations Not Used           0


                                        Iteration History

                   Iteration    Evaluations    -2 Res Log Like       Criterion

                           0              1       216.06796912
                           1              2       205.11045832      0.00000263
                           2              1       205.11027324      0.00000000


                                    Convergence criteria met.
```

```
                                       The Mixed Procedure

                         Estimated R Correlation Matrix for Subject_ 10002

            Row        Col1        Col2        Col3        Col4        Col5        Col6

              1      1.0000      0.5394      0.2910      0.1570     0.08468     0.04568
              2      0.5394      1.0000      0.5394      0.2910      0.1570     0.08468
              3      0.2910      0.5394      1.0000      0.5394      0.2910      0.1570
              4      0.1570      0.2910      0.5394      1.0000      0.5394      0.2910
              5     0.08468      0.1570      0.2910      0.5394      1.0000      0.5394
              6     0.04568     0.08468      0.1570      0.2910      0.5394      1.0000


                                 Covariance Parameter Estimates

                                Cov Parm     Subject     Estimate

                                AR(1)        Subject_      0.5394
                                Residual                  16.1927


                                         Fit Statistics

                              -2 Res Log Likelihood           205.1
                              AIC (Smaller is Better)         209.1
                              AICC (Smaller is Better)        209.5
                              BIC (Smaller is Better)         209.0


                                 Null Model Likelihood Ratio Test

                                   DF    Chi-Square      Pr > ChiSq

                                    1         10.96          0.0009


                                  Type 3 Tests of Fixed Effects

                                            Num     Den
                     Effect                  DF      DF    F Value    Pr > F

                     visit_num                5      30       1.11    0.3769
                     CigarettePerday_bl       1       5      31.69    0.0025
```

Questionnaire of Smoking Urges-Brief (QSU-B)

Note: The “QSUdata2” dataset is in long format, with a baseline measure and 2 measurements during each of 5 rTMS sessions (1 pre-session and 1 post-session).

/* full model – the AIC from this model was 408.3, indicating that this model with a compound symmetry error structure provided a better fit than the model with an AR(1) structure, which had an AIC of 416.4 */

proc mixed data=QSUdata2;

class subjectID visitNumber TrGroup prepost;

model qsu_score = TrGroup visitNumber TrGroup*visitNumber prepost TrGroup*prepost visitNumber*prepost TrGroup*visitNumber*prepost qsu_baseline;

repeated / subject=subjectID type=cs rcorr;

run;

/* null model - for aid in calculation of f squared statistic */

proc mixed data=QSUdata2;

class subjectID visitNumber TrGroup prepost;

model qsu_score = visitNumber prepost qsu_baseline;

repeated / subject=subjectID type=cs rcorr;

run;

Output from model estimation for the “full model”:

```
                                       The Mixed Procedure

                                        Model Information

                      Data Set                     WORK.QSUDATA2
                      Dependent Variable           qsu_score
                      Covariance Structure         Compound Symmetry
                      Subject Effect               subjectID
                      Estimation Method            REML
                      Residual Variance Method     Profile
                      Fixed Effects SE Method      Model-Based
                      Degrees of Freedom Method    Between-Within


                                     Class Level Information

                     Class          Levels    Values

                     subjectID           7    10002 10003 10004 10005 20001
                                              20002 20003
                     visitNumber         5    1.00 2.00 3.00 4.00 5.00
                     TrGroup             2    X Y
                     PrePost             2    POST PRE


                                           Dimensions

                               Covariance Parameters             2
                               Columns in X                     55
                               Columns in Z                      0
                               Subjects                          7
                               Max Obs per Subject              10


                                     Number of Observations

                           Number of Observations Read              70
                           Number of Observations Used              70
                           Number of Observations Not Used           0


                                        Iteration History

                   Iteration    Evaluations    -2 Res Log Like       Criterion

                           0              1       422.20410376
                           1              1       404.34306758      0.00000000


                                    Convergence criteria met.


                        Estimated R Correlation Matrix for subjectID 10002

   Row      Col1      Col2      Col3      Col4      Col5      Col6      Col7      Col8      Col9

     1    1.0000    0.4749    0.4749    0.4749    0.4749    0.4749    0.4749    0.4749    0.4749
     2    0.4749    1.0000    0.4749    0.4749    0.4749    0.4749    0.4749    0.4749    0.4749
     3    0.4749    0.4749    1.0000    0.4749    0.4749    0.4749    0.4749    0.4749    0.4749
     4    0.4749    0.4749    0.4749    1.0000    0.4749    0.4749    0.4749    0.4749    0.4749
     5    0.4749    0.4749    0.4749    0.4749    1.0000    0.4749    0.4749    0.4749    0.4749
     6    0.4749    0.4749    0.4749    0.4749    0.4749    1.0000    0.4749    0.4749    0.4749
     7    0.4749    0.4749    0.4749    0.4749    0.4749    0.4749    1.0000    0.4749    0.4749
     8    0.4749    0.4749    0.4749    0.4749    0.4749    0.4749    0.4749    1.0000    0.4749
     9    0.4749    0.4749    0.4749    0.4749    0.4749    0.4749    0.4749    0.4749    1.0000
    10    0.4749    0.4749    0.4749    0.4749    0.4749    0.4749    0.4749    0.4749    0.4749

                                            Estimated R
                                            Correlation
                                            Matrix for
                                             subjectID
                                               10002

                                           Row     Col10

                                             1    0.4749
                                             2    0.4749
                                             3    0.4749
                                             4    0.4749
                                             5    0.4749
                                             6    0.4749
                                             7    0.4749
                                             8    0.4749
                                             9    0.4749
                                            10    1.0000


                                  Covariance Parameter Estimates

                                Cov Parm     Subject      Estimate

                                CS           subjectID     80.6655
                                Residual                   89.2085


                                         Fit Statistics

                              -2 Res Log Likelihood           404.3
                              AIC (Smaller is Better)         408.3
                              AICC (Smaller is Better)        408.6
                              BIC (Smaller is Better)         408.2


                                 Null Model Likelihood Ratio Test

                                   DF    Chi-Square      Pr > ChiSq

                                    1         17.86          <.0001


                                  Type 3 Tests of Fixed Effects

                                             Num     Den
                    Effect                    DF      DF    F Value    Pr > F

                    TrGroup                    1       4       0.01    0.9382
                    visitNumber                4      20       3.99    0.0154
                    visitNumber*TrGroup        4      20       6.96    0.0011
                    PrePost                    1       5       0.39    0.5601
                    TrGroup*PrePost            1       5       2.44    0.1791
                    visitNumber*PrePost        4      20       0.88    0.4910
                    visitN*TrGrou*PrePos       4      20       0.80    0.5379
                    qsu_baseline               1       4      61.62    0.0014
```

Output from model estimation for the “null model”:

```
                                       The Mixed Procedure

                                        Model Information

                      Data Set                     WORK.QSUDATA2
                      Dependent Variable           qsu_score
                      Covariance Structure         Compound Symmetry
                      Subject Effect               subjectID
                      Estimation Method            REML
                      Residual Variance Method     Profile
                      Fixed Effects SE Method      Model-Based
                      Degrees of Freedom Method    Between-Within


                                     Class Level Information

                     Class          Levels    Values

                     subjectID           7    10002 10003 10004 10005 20001
                                              20002 20003
                     visitNumber         5    1.00 2.00 3.00 4.00 5.00
                     TrGroup             2    X Y
                     PrePost             2    POST PRE


                                           Dimensions

                               Covariance Parameters             2
                               Columns in X                      9
                               Columns in Z                      0
                               Subjects                          7
                               Max Obs per Subject              10


                                     Number of Observations

                           Number of Observations Read              70
                           Number of Observations Used              70
                           Number of Observations Not Used           0


                                        Iteration History

                   Iteration    Evaluations    -2 Res Log Like       Criterion

                           0              1       530.41496981
                           1              1       519.05777415      0.00000000


                                    Convergence criteria met.


                        Estimated R Correlation Matrix for subjectID 10002

   Row      Col1      Col2      Col3      Col4      Col5      Col6      Col7      Col8      Col9

     1    1.0000    0.3212    0.3212    0.3212    0.3212    0.3212    0.3212    0.3212    0.3212
     2    0.3212    1.0000    0.3212    0.3212    0.3212    0.3212    0.3212    0.3212    0.3212
     3    0.3212    0.3212    1.0000    0.3212    0.3212    0.3212    0.3212    0.3212    0.3212
     4    0.3212    0.3212    0.3212    1.0000    0.3212    0.3212    0.3212    0.3212    0.3212
     5    0.3212    0.3212    0.3212    0.3212    1.0000    0.3212    0.3212    0.3212    0.3212
     6    0.3212    0.3212    0.3212    0.3212    0.3212    1.0000    0.3212    0.3212    0.3212
     7    0.3212    0.3212    0.3212    0.3212    0.3212    0.3212    1.0000    0.3212    0.3212
     8    0.3212    0.3212    0.3212    0.3212    0.3212    0.3212    0.3212    1.0000    0.3212
     9    0.3212    0.3212    0.3212    0.3212    0.3212    0.3212    0.3212    0.3212    1.0000
    10    0.3212    0.3212    0.3212    0.3212    0.3212    0.3212    0.3212    0.3212    0.3212

                                            Estimated R
                                            Correlation
                                            Matrix for
                                             subjectID
                                               10002

                                           Row     Col10

                                             1    0.3212
                                             2    0.3212
                                             3    0.3212
                                             4    0.3212
                                             5    0.3212
                                             6    0.3212
                                             7    0.3212
                                             8    0.3212
                                             9    0.3212
                                            10    1.0000


                                  Covariance Parameter Estimates

                                Cov Parm     Subject      Estimate

                                CS           subjectID     59.2661
                                Residual                    125.25


                                         Fit Statistics

                              -2 Res Log Likelihood           519.1
                              AIC (Smaller is Better)         523.1
                              AICC (Smaller is Better)        523.3
                              BIC (Smaller is Better)         522.9


                                 Null Model Likelihood Ratio Test

                                   DF    Chi-Square      Pr > ChiSq

                                    1         11.36          0.0008


                                  Type 3 Tests of Fixed Effects

                                         Num     Den
                        Effect            DF      DF    F Value    Pr > F

                        visitNumber        4      24       2.02    0.1243
                        PrePost            1       6       0.52    0.4974
                        qsu_baseline       1       5      78.78    0.0003
```

Carbon Monoxide Levels

Note: The “CO2” dataset is in long format, with a carbon monoxide measurement during each of 5 rTMS sessions.

SAS programming code:

/* full model – note that the model using an AR(1) covariance structure did not converge and thus the compound symmetry structure was used. */

proc mixed data=co2;

class subjectidco visit_num TrGroupco ;

model co = TrGroupco visit_num TrGroupco*visit_num co_bl;

repeated int / subject=subjectidco type=cs rcorr;

run;

/* null model - for aid in calculation of f squared statistic */

proc mixed data=co2 ;

class subjectidco visit_num ;

model co = visit_num co_bl;

repeated int / subject=subjectidco type=cs r;

run;

Output from model estimation for the “full model”:

```
                                       The Mixed Procedure

                                        Model Information

                      Data Set                     WORK.CO2
                      Dependent Variable           CO
                      Covariance Structure         Compound Symmetry
                      Subject Effect               subjectIDCO
                      Estimation Method            REML
                      Residual Variance Method     Profile
                      Fixed Effects SE Method      Model-Based
                      Degrees of Freedom Method    Between-Within


                                     Class Level Information

                     Class          Levels    Values

                     subjectIDCO         7    10002 10003 10004 10005 20001
                                              20002 20003
                     visit_num           5    TMS1 TMS2 TMS3 TMS4 TMS5
                     TrGroupCO           2    active sham


                                           Dimensions

                               Covariance Parameters             2
                               Columns in X                     19
                               Columns in Z                      0
                               Subjects                          7
                               Max Obs per Subject               5


                                     Number of Observations

                           Number of Observations Read              35
                           Number of Observations Used              35
                           Number of Observations Not Used           0


                                        Iteration History

                   Iteration    Evaluations    -2 Res Log Like       Criterion

                           0              1       137.66424417
                           1              1       136.37047204      0.00000000


                                    Convergence criteria met.


                       Estimated R Correlation Matrix for subjectIDCO 10002

                  Row        Col1        Col2        Col3        Col4        Col5

                    1      1.0000      0.1996      0.1996      0.1996      0.1996
                    2      0.1996      1.0000      0.1996      0.1996      0.1996
                    3      0.1996      0.1996      1.0000      0.1996      0.1996
                    4      0.1996      0.1996      0.1996      1.0000      0.1996
                    5      0.1996      0.1996      0.1996      0.1996      1.0000


                                  Covariance Parameter Estimates

                               Cov Parm     Subject        Estimate

                               CS           subjectIDCO      1.6533
                               Residual                      6.6283


                                         Fit Statistics

                              -2 Res Log Likelihood           136.4
                              AIC (Smaller is Better)         140.4
                              AICC (Smaller is Better)        140.9
                              BIC (Smaller is Better)         140.3


                                 Null Model Likelihood Ratio Test

                                   DF    Chi-Square      Pr > ChiSq

                                    1          1.29          0.2554


                                  Type 3 Tests of Fixed Effects

                                             Num     Den
                     Effect                   DF      DF    F Value    Pr > F

                     TrGroupCO                 1       4       0.27    0.6300
                     visit_num                 4      20       1.04    0.4128
                     visit_num*TrGroupCO       4      20       1.14    0.3680
                     co_bl                     1       4     111.07    0.0005
```

Output from model estimation for the “null model”:

```
                                                                                 The SAS System     08:43 Wednesday, January 26, 2022  59

                                       The Mixed Procedure

                                        Model Information

                      Data Set                     WORK.CO2
                      Dependent Variable           CO
                      Covariance Structure         Compound Symmetry
                      Subject Effect               subjectIDCO
                      Estimation Method            REML
                      Residual Variance Method     Profile
                      Fixed Effects SE Method      Model-Based
                      Degrees of Freedom Method    Between-Within


                                     Class Level Information

                     Class          Levels    Values

                     subjectIDCO         7    10002 10003 10004 10005 20001
                                              20002 20003
                     visit_num           5    TMS1 TMS2 TMS3 TMS4 TMS5


                                           Dimensions

                               Covariance Parameters             2
                               Columns in X                      7
                               Columns in Z                      0
                               Subjects                          7
                               Max Obs per Subject               5


                                     Number of Observations

                           Number of Observations Read              35
                           Number of Observations Used              35
                           Number of Observations Not Used           0


                                        Iteration History

                   Iteration    Evaluations    -2 Res Log Like       Criterion

                           0              1       158.93382303
                           1              1       157.99852770      0.00000000


                                    Convergence criteria met.


                       Estimated R Correlation Matrix for subjectIDCO 10002

                  Row        Col1        Col2        Col3        Col4        Col5

                    1      1.0000      0.1492      0.1492      0.1492      0.1492
                    2      0.1492      1.0000      0.1492      0.1492      0.1492
                    3      0.1492      0.1492      1.0000      0.1492      0.1492
                    4      0.1492      0.1492      0.1492      1.0000      0.1492
                    5      0.1492      0.1492      0.1492      0.1492      1.0000


                                  Covariance Parameter Estimates

                               Cov Parm     Subject        Estimate

                               CS           subjectIDCO      1.1891
                               Residual                      6.7786


                                         Fit Statistics

                              -2 Res Log Likelihood           158.0
                              AIC (Smaller is Better)         162.0
                              AICC (Smaller is Better)        162.5
                              BIC (Smaller is Better)         161.9


                                 Null Model Likelihood Ratio Test

                                   DF    Chi-Square      Pr > ChiSq

                                    1          0.94          0.3335


                                  Type 3 Tests of Fixed Effects

                                        Num     Den
                          Effect         DF      DF    F Value    Pr > F

                          visit_num       4      24       1.11    0.3745
                          co_bl           1       5     141.40    <.0001
```
